# The Governance of Pandemics in Primary Health Care: The Governance Strategies Adopted by Health Facility Governing Committees in Times of COVID Pandemic in Tanzania

**DOI:** 10.1101/2022.01.03.22268663

**Authors:** Anosisye M. Kesale, Mikidadi Muhanga, Christopher Mahonge

## Abstract

The governance of COVID 19 in Lower and Middle-Income Countries (LMICs) is very critical for curbing its effects. However, it is unknown what governance strategies are adopted by Health Facility Governing Committees (HFGCs) s as a response to the pandemic. We employed an exploratory qualitative design to study the governance strategies adopted by HFGCs during the COVID19. Since COVID 19 is new, an inductive approach was used as it involves analyzing collected data with little or no predetermined theory for the study. A purposive sampling technique was employed in which multistage clustered sampling was used to select regions, councils, health facilities and respondents. In-depth interviews with HFGCs chairpersons and Focus Group Discussions with members of HFGCs were used to collect data. The data were analyzed based on the themes which emerged during data collection. We found five governance strategies that were found to be commonly adopted by many HFGCs which are financial allocation, re-planing, mobilization of resources, community sensitization and mobilization of stakeholders. however, these governance structures were not all adopted by all HFGCs. The HFGCs slowly adopted governance strategies in the times of COVID 19 pandemics because were unprepared. Despite being empowered by the Direct Health Facility Financing, still, the newest of the COVID 19 has been a challenge to many HFGCs. This calls for urgent capacity building for governance institutions on how to deal will pandemics in primary health facilities.

**Key Questions Box:** **Key Questions**

What is Already Known?

➢ Governance of pandemics is very critical for minimizing its effects
➢ Government in Lower and Middle-Income Countries (LMICs) reacted differently to bring the COVID 19 pandemic under control
➢ Little is known on the governance strategies adopted by Health Facility Governing Committees (HFGCs) to control COVID 19 at the primary health care facilities in LMICs

What are the new findings?

➢ HFGCs response to the COVID 19 is mixed as some HFGCs were so active and others were so slowly in instituting governance strategies to combat the pandemic
➢ The common governance strategies adopted were financial allocation, re-planning, embolization of resources, community sensitization and stakeholders mobilization
➢ HFGCs in LMICs were not well prepared for the pandemic at the community level

What do new findings imply?

➢ Capacitated HFGCs in both soft and hard aspects cornerstone for effective governance of the pandemics in primary health care
➢ Preparedness of pandemics in LMICs needs to go beyond upper-level governance structures

## Introduction

Governance is appreciated to be the foundation for improving health service delivery at the primary health care (PHC) [1]. It is indeed critical for achieving Universal Health Coverage (UHC) and Sustainable Development Goal (SDG) number 6. Therefore government’s efforts to strengthen governance at all levels promise better health outcomes and improved wellbeing of the population [2]. The benefits of governance can be achieved through having effective, accountable and more inclusive governance institutions. Lower and Middle-Income Countries (LMICs) have applied various policies to strengthen governance at PHC. The common policy that is being applied in LMICs is decentralization through various types such as fiscal, administrative and political decentralization [3], [4]. Decentralization policy allows the transfer of administrative, fiscal and decision-making powers and responsibilities to the subnational governing entity for overseeing health service delivery. At the primary health care facilities, decentralization has created community governance structures known as Health Facility Governing Committees (HFGCs) to govern health service delivery [5]. These HFGCs are composed of local users responsible for monitoring health service providers, improving local choices and participation of communities in influencing the quality, responsiveness and coverage of health service[6].

The governance of pandemics is very critical for curbing and responding to several infectious pandemics [7]. Under the COVID-19 Pandemic, all governance levels including primary health care are tested, therefore, calls for new governance strategies to be taken by governance institutions [8]. The literature ascertain that the pandemics are characterized by unclear definition of problems, inconsistency, lack of standards solution and have conflicting goals and cultures, therefore, requires functional and innovative governance institutions to address those problems[8]–[11]. Therefore, traditional governance strategies can no longer be effective in solving pandemics’ problems because of their nature. In turbulent or critical situations such as COVID-19, governance institutions are required to adopt creative and agile strategies [12], [13]. This will help to protect health facilities, their staff and the population through establishing networks, partnerships with stakeholders, mobilizing resources and re-establishing new plans [1], [7], [8], [14]. Therefore, effective governance of pandemics requires effective governance mechanisms need to be decided and taken by the existing governance institutions [1], [15].

In LMICs the governance of primary health care facilities is decentralized to community governance structures through HFGCs[16]. Through this people’s centered governance approach, health service users are devolved with powers and mandates to oversee and strengthen service utilization, responsiveness and increase providers’ accountability in times of COVID-19 pandemic [6]. Like any other pandemics, COVID-19 tends to undermine and disturb the pre-determined health facility plans and budgets due to its unprecedented characteristics [12], [17]. To address challenges posed by COVID-19 HFGCs need to make divergent and timely decisions to enhance facility operations and protect community health. Since the challenges posed by the governance of COVID-19 are not only technical, the governance of COVID-19 should adopt a mix of socio-political and cultural strategies in which HFGCs innovation and creativity is very key element [12], [14], [18]. The adequate governance of primary health care facilities practiced by HFGCs determines the health outcomes of a particular community. This is because HFGCs are responsible for making all major decisions regarding health facilities operations and indeed link the communities and service providers [17], [19], [20].

In a Tanzanian decentralized health care system, HFGCs were introduced in 1999 during the health sector reforms [21]. The HFGCs are composed of elected community members and health facilities in charge [22]. These HFGCs are assigned specific governance functions to perform such as participating in the planning, budgeting and procurement process. Mobilizing people to join community health funds and collecting, discussing and addressing community health challenges. Furthermore, HFGCs are responsible for participating in monitoring renovation and construction activities of the facility and working with different stakeholders and partners to mobilize resources for the health facilities [23]. The government of Tanzania has continued to embark on different reforms to strengthen the health system including empowering HFGCs to accomplish their devolved functions. The current reform pursued by the government of Tanzania is Direct Health Facility Financing (DHFF). The Government of Tanzania (GoT) decided to introduce Direct Health Facility Financing (DHFF) to ensure flexible and timely funding and use at the level of service delivery points to ensure increased efficiency in financial use, accountability and quality service delivery to the public [24], [25]. The DHFF initiative further aligns with global health initiatives such as the Universal Health Coverage (UHC) and Sustainable Development Goals (SDGs) [26]. Despite this massive reform in the primary health care facilities, it is not known how and what governance strategies were adopted by the empowered HFGCs in primary health care to govern the COVID 19 pandemic Tanzania. This study was conducted to assess the governance initiatives instituted by the HFGCs in selected primary health facilities in Tanzania.

## Methods

### Method and Approach

We employed a qualitative method to understand the governance of pandemics in primary health care facilities in two selected regions (Kilimanjaro and Songwe) in Tanzania between February to April 2021. Kilimanjaro region was selected because is among the region with high health facilities and HFGCs performance as per the Star Rating Assessment conducted by the Presidents Office-Regional Administration and Local Government of Tanzania in all primary health facilities 2018 [27]. Songwe region was selected because is among the region with low health facilities and HFGCs performance as per the star rating assessment conducted in 2018. An inductive approach was used because it involves analyzing collected data with little or no predetermined theory for the study. This is because the governance of COVID-19 is a new phenomenon, therefore, no theory explains how COVID-19 should be governed. Therefore the actual data collected from the sites determined the analysis of the data. An inductive approach was adopted despite being time-consuming because it offers a lot where the phenomenon studied is new and not known.

### Study Design

This study employed an exploratory qualitative design to study the governance strategies adopted by HFGCs during the COVID-19 pandemic. The selected design was deemed appropriate as the engaging community through the HFGCs is not a linear process as it involves social processes. Since the COVID-19 pandemic is a new disease and the roles of HFGCs have not been documented, a flexible design was necessary to lay the foundation for other studies by generating new insight into the phenomenon.

### Sampling Technique and Sample Size

A purposive sampling technique was employed to select regions, councils and respondents to the study. Regions and councils were selected based on their performance as per the star rating assessment conducted in 2018 in all primary health facilities in Tanzania. after the selection of the two regions Kilimanjaro with high performing facilities and HFGCs but also found in the northern part of Tanzania, Songwe was selected because was the region with low performing health facilities and HFGCs but also found in the southern highland of the country. Two councils were then selected from each region, some factors determined their selection. In Kilimanjaro, Moshi Municipal was selected because was the council with low health facilities and HFGCs performance but also in the urban local authority. Siha district council was selected because is the top performer countrywide as per the star rating assessment conducted in 2018 but also is the rural local authority. In Songwe likewise, Tunduma Town council was selected because has high performance in the region but also is an urban authority bordered to Zambia, therefore, providing uniqueness to our sample. Mbozi district council was selected because is the lower performer in the region but also a district council, indeed Mbozi is a headquarter of the Songwe region. From each council, two high-performing health facilities and two low-performing primary health facilities were selected. Before the selection, the health facilities were grouped into two categories the health centers and dispensaries. Therefore from each council, a high-performing health center and dispensary were selected. Also, a low-performing health center and dispensary were purposively selected. From each selected health facility, respondents were drawn. The selection criteria were based on the ability of the respondents to provide relevant information about the governance of health facilities during COVID-19. Therefore, all members of HFGCs were selected for FGDs and only HFGCs Chairpersons of the selected primary health facilities implementing DHFF qualified for interviews. A total number of 14 interviews were conducted until the saturation point and FGDs were conducted to the saturation point. Saturation was reached when the participant started to provide a similar response and no new information was added. Each FGD consisted of 6 to 9 participants who are members of the HFGCs. The highest number of participants was 9 because the total number of HFGC members is 9 for health centers and 8 for dispensaries.

### Data Collection methods

Data collection for this study was conducted between February and April 2021. This was the period when Tanzania and the world, in general, experienced the second wave of COVID-19. The study employed in-depth interviews and Focus Group Discussions to collect data from the respondents. In-depth interviews were used to collect data from the HFGCs chairpersons while the FGDs were employed to collect data from all HFGC members. interview and FGDs guide were developed based on the main area of our focus which aimed at understanding the governance strategies adopted by the HFGCs during the COVID-19 pandemics. Then from this main area of our focus sub-topics of what to be discussed were predetermined to understand the experience of HFGCs members on dealing with the pandemic. respondents were given the freedom to express their experience with some probing questions to get more details on the way they participated in the COVID-19 pandemic.

### Data Analysis

The collected data of this study were analyzed based on the themes which emerged during data collection. The in-depth interviews and FGDs conducted were audio recorded. The audio recorded was transcribed verbatim. This allowed researchers to read and re-read the extracts. The written extracts were then reduced into codes that were informed by the focus areas of the study that was the governance strategies adopted by HFGCs in times of COVID-19 pandemic. Therefore all specific areas in which respondents touched the issue of the governance of COVID-19 were coded. Significant themes relating to governance strategies induced by HFGCs in the times of the COVID-19 pandemic were captured from the generated codes, the generated themes were linked to the focus area of the study. Five themes from the codes were generated as indicated in table 1. All the governance themes generated were reviewed and enriched from the collected data to be more meaningful and correlate with the study topic and the overlapping themes were merged into one theme. The themes were then refined and defined well to give meaning to the reader on what they imply in the governance of the COVID-19 pandemic context.

**Table 1:**
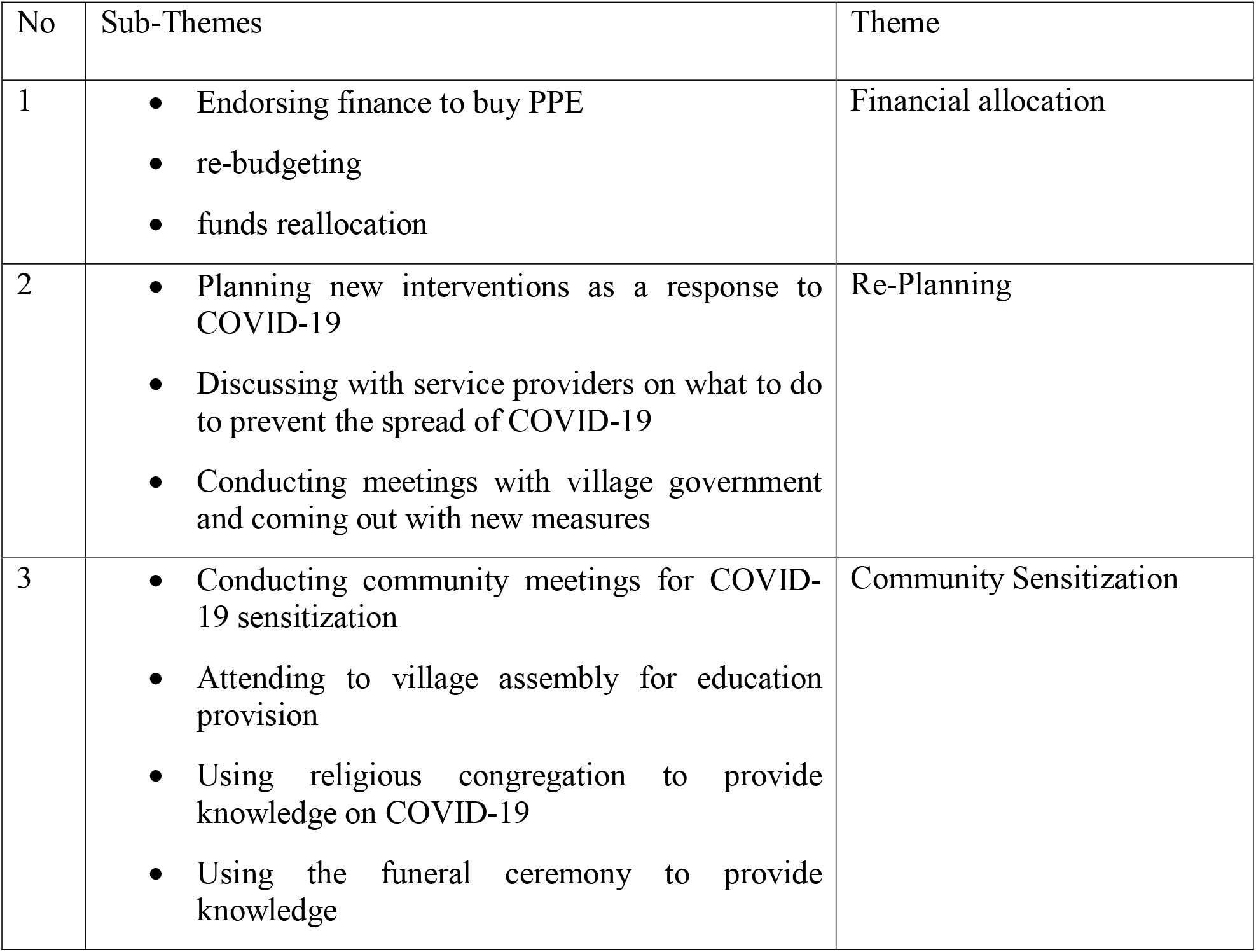
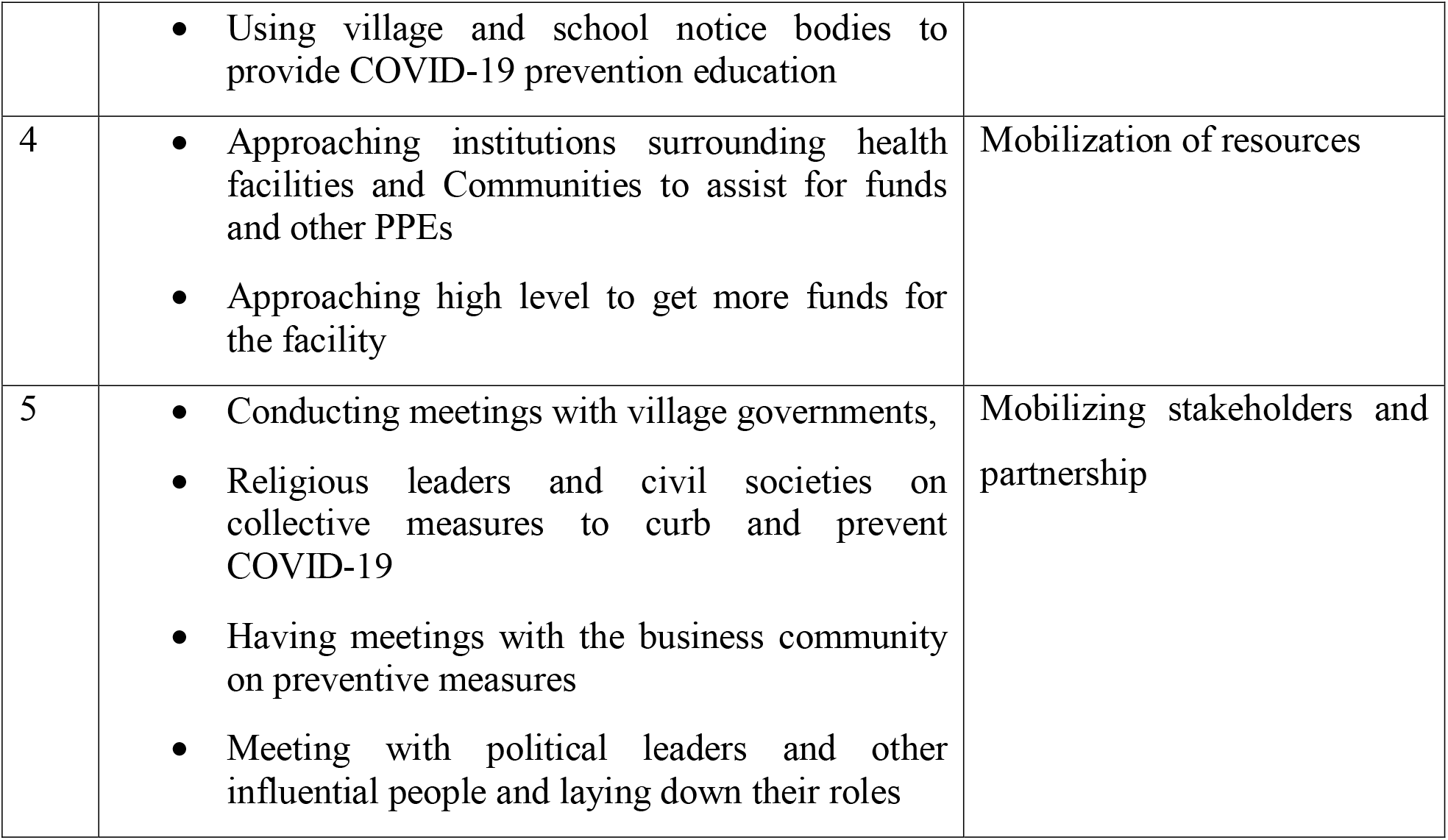
A summary of themes and sub-themes that emerged in the data collection

| No | Sub-Themes | Theme |
| --- | --- | --- |
| 1 | • Endorsing finance to buy PPE<br>• re-budgeting<br>• funds reallocation | Financial allocation |
| 2 | • Planning new interventions as a response to COVID-19<br>• Discussing with service providers on what to do to prevent the spread of COVID-19<br>• Conducting meetings with village government and coming out with new measures | Re-Planning |
| 3 | • Conducting community meetings for COVID-19 sensitization<br>• Attending to village assembly for education provision<br>• Using religious congregation to provide knowledge on COVID-19<br>• Using the funeral ceremony to provide knowledge | Community Sensitization |
|  | • Using village and school notice bodies to provide COVID-19 prevention education |  |
| 4 | • Approaching institutions surrounding health facilities and Communities to assist for funds and other PPEs<br>• Approaching high level to get more funds for the facility | Mobilization of resources |
| 5 | • Conducting meetings with village governments,<br>• Religious leaders and civil societies on collective measures to curb and prevent COVID-19<br>• Having meetings with the business community on preventive measures<br>• Meeting with political leaders and other influential people and laying down their roles | Mobilizing stakeholders and partnership |

### Ethical Declaration

This study was conducted in accordance with the principles of the Declaration of Helsinki. All methods were carried out in accordance with relevant guidelines and regulations. The ethical clearance was sought from the Sokoine University of Agriculture and granted by the National Institute of Medical Research (NIMR)- (Ref.No.NIMR/HQ/R.8a/Vol.IX/2740. The permit was then submitted to the President’s Office Regional Administration and Local Government (PO-RALG) to be permitted to research local government authorities. PO-RALG offered a permit with registration number AB.307/323/01 to allow the research to research the selected regions. Informed consent was obtained from all human participants of this study by completing the consent forms before they were involved in the study

## Results

Five Themes were generated from the collected data namely Community sensitization, financial allocation, Re-planning, resource mobilization and mobilization of stakeholders and partnerships. The themes emerged from sub-themes which came out after asking the participants what governance strategies they employed as a response to COVID-19 in their primary health facilities.

Themes and sub-themes are presented in detail below together with interview quotes that aim at supporting and providing illustration to the presented concepts.

### Community Sensition on COVID-19

This theme portrays participants perception regarding the HFGCs governance decision they made about the community sensitization on COVID-19

Participants agree that at least they took some measures including providing education about the COVID-19 and sensitized the community on how to prevent themselves from contracting the pandemic. Participants mentioned some of the strategies they used to sensitize the community which are having a special meeting with communities and educating about the pandemic, attending other meetings which were conducted within their village or around their health facilities. Other strategies employed were some members of HFGCs were attending religious congregation and were given time to educate the community and also using funeral ceremony for the same. For instance one of the HFGC Chairperson had this to say.

> *“We identified some strategies to reach the community and build awareness on the COVID-19… then our members were spread all over the village in every Sunday to provide awareness through churches”*(HFGC Chairperson-Siha District Council)

Others had this to say

> *“We did not involve ourselves in providing education to the community because even us were not much aware of the pandemic but also in our village we restricted all mass meetings including religious activities”* (HFGCs Chairperson-Mbozi District Council)

### Re-Planing

The theme and sub-themes portrays the governance initiatives taken by the HFGCs regarding the planning of health interventions to combat COVID-19

The participants reported that since the pandemic was new and never foreseen during the health facility plan preparation, they had to revisit their existing plan and come up with new plans or interventions which could help the facility to respond to the COVID-19. Participants agreed that they used special and normal meetings to generate new strategies to be implemented by the health facility. They reported that they had to invite some village government leaders and civil societies to discuss important interventions to be implemented by the facilities and community such as buying PPE which was not a part of the facility plans. Also, some of the washing hands’ pieces of equipment were procured by the facility to enable patients who attended the facility to wash their hands. Participants of FGDs had this to say

> *“As soon as we got information about the pandemic we advised our HFGC chairperson to prepare a meeting which involved a facility staff and HFGC… then in that meeting, they educated us on the pandemic and advised us what should be done. We together we came out with new plans which were to be implemented by our facilities and others which were later communicated to the community”*(HFGC Chairperson-Siha)

However, some other participants had a different opinion on this, as they responded that the facility did not revisit their plan rather than seeing health providers buying some of the equipment for the health facility.

> *“Our committee did nothing for sure on this*.. *but our facility in charge procured everything…he just asked us to endorse funds to procure medicine and medical equipment”* (HFGC Chairperson-Mbozi)

### Financial Allocation

The present themes express the participants experience on the HFGCs governance initiatives made to fund COVID-19 related activities

Participants were on the view that the rise of the COVID-19 forced them to go back to the drawing body, as they had to do some reallocation of funds that were budgeted for other interventions. They agreed that since currently, they implementing activities-based budgeting, so they were advised by the facility staff to endorse some reallocation to help to implementation of the new COVID-19 interventions that were planed.

> *“Through our HFGC meetings were endorsed reallocation of funds which originally were budgeted for the renovation of the health facility…. Therefore, part of the fund had now to be used to fund COVID-19 issues”* (HFGC Chairperson-Moshi Municipal Council)

Other responded

> *“So far no financial decision has been made by our HFGC because COVID-19 was not part of our facility plan. also, we are hesitant to plan anything which will include finance because we haven’t received our funds from the central government and other sources like health insurance”* (HFGC chairperson-Mbozi)

### Resource Mobilization

The present theme describes the HFGCs experience on the governance initiatives regarding

### Resource mobilization

Participants through both in-depth interviews and FGDs reacted that as a governance entity they were involved in soliciting resources from their stakeholders and partners. They revealed that they identified facility and community needs and presented them to their stakeholders for help. They agreed that some stakeholders assisted them with funds while some others contributed equipment such as water tanks, masks, sanitizers while others did not respond at all.

> *“We wrote letters and submitted to our stakeholders whom we thought were able to help us… the response was superb as we managed to secure some resources which were very useful”(*HFGC Chairperson-Tunduma)

Other said

> *“We did nothing for sure because we’re not aware that our HFGC was responsible for all that “* (HFGC Chairperson-Moshi)

### Mobilizing stakeholders and establishing partnerships

This theme presents the participant’s perception of the HFGCs governance initiatives taken relating to the mobilization of resources and establishing partnerships

Participants in several scenarios agreed that they mobilized all health-related stakeholders such as civil societies, the business community, political leaders and faith-based institutions to come together to fight for the pandemic. Some members agree that stakeholders played a big part because they offered resources which facility could not manage to offer to the community. Given the scarcity of a resource in primary health facilities, the strategy helped to get extra resources but established new partnerships with the facility and HFGCs.

> *“Look*.. *stakeholders were happy to work with us because we engaged and we were very transparent…. They donated so many things including providing fuel to our ambulance*” (HFGC Chairperson-Tunduma)

Some other participants had a different opinion regarding the mobilization of the stakeholders and partners during the pandemic. They believed that they did not do anything concerning meeting or establishing any partnership with an outsider to help to curb COVID-19. Through the FGDs one of the respondents responded

> *“I think it not within our mandates to meet with stakeholders, maybe health workers can do that…. For us here we never engaged in that aspects”* (HFGC Chairperson-Siha)

The above explanation and quotation from both in-depth interviews and FGDs have been summarized in Table 1.

## Discussion

In the governing pandemics such as COVID-19 in primary health care, more can be done by the governance structures to respond and curb the COVID-19. There is a mixed experience of HFGCs regarding the governance initiatives taken in time of the COVID-19 pandemic. Some HFGCs were so active in using their devolved governance powers and mandates to make decisions to combat COVID-19 while some other HFGCs did not manage to make governance decisions as was expected. However, this study has identified the common governance strategies adopted by HFGCs in selected primary health facilities in Tanzania in the context of DHFF. the study has found that HFGCs adopted governance strategies such as budgeting allocation for COVID-19 related activities, re-planing to accommodate COVID activities, community sensitization, mobilization of stakeholders and resource mobilization. These strategies were found to be significant for setting the direction of the health facilities and communities during the COVID-19. The governance strategies were found to be adopted by the majority of the HFGCs differently. The adopted strategies were found to be within the devolved HFGCs governance responsibilities and powers.

Participants responded that the HFGCs and primary health facilities were slow in responding to the COVID-19 because they were not prepared for the pandemic therefore they were not aware of the appropriate strategies which would be appropriate for combating the pandemic. Shortage of resources such as finance also was found to be the major impediment that influenced HFGC’s dilemma to adopt some of the governance strategies. This result is in line with the arguments raised by some literature that health system unpreparedness during the rise of COVID-19 was predicted to negatively impact the fight against the pandemic [28]–[30]. Despite all the setbacks, HFGCs to the large extent played their part by ensuring the availability of basic things health facilities such as PPE, masks and sanitizers. Through the mobilization of stakeholders and resources that governance initiatives helped to fill the gap of shortage of funding in primary health facilities as medical commodities and medicines required were obtained. This is also in line with [15]who stated the importance of collective efforts among health stakeholders in dealing with the COVID 19.

Despite recommendable governance strategies adopted by most of HFGCs in the course of governing primary health care facilities as found by this study, still many respondents have indicated unpreparedness of the governance institutions from the national level to the subnational level such as HFGCs in governing the pandemics. This is because HFGCs have been found to have struggled a lot to make decisions that are within their ambits. In some health facilities, HFGCs were reluctant to make decisions that are part of their functions because they were not aware of that. This was predicted by [30] that delay in deciding and instituting appropriate measures by government entities and health managers would put more risk to the lives and death of community members and health workers. We argue that HFGCs and other governance institutions can only make appropriate decisions if they are prepared enough on how they should use and accomplish their devolved powers and responsibilities [31]–[33]. More exciting, before the implementation of DHFF, HFGCs had limited powers to initiate any intervention to be implemented in the health facilities because such powers were vested to the Council level and primary health care level. However, after the decentralization of fiscal and decision-making powers through DHFF to the primary health care facilities, some of the HFGCs are found to be enjoying their mandates and powers through initiating several interventions to protect their communities and health workers. However, other HFGCs are not yet to use such powers and mandates as has been revealed in this study.

From the result of this study, governance of the pandemics by the HFGCs at the primary health care facilities has been based on the specific functions devolved to HFGCs. this experience is in line with the main of Alma Ata declaration of 1978 that insisted community participation through community governance structures to monitor health service delivery [34]–[36]. In the COVID 19 context, HFGCs have played their part to some extent as was expected though there is a long way to go. Indeed, the DHFF arrangement has empowered HFGCs to come out with several interventions such as financial allocation and planning that could not be possible initiated prior to DHFF implementation [5], [37]. As suggested by [1], [38] frequently strengthening governance institutions are fundamental for proving relevant governance decisions in times of pandemics. This is the impact of DHFFin the decisions that have been made by some of the health facilities in addressing the COVID 19 pandemic.

COVID 19 being a novel disease in the World and so in LMICs, it has caused many effects to the health workers, communities and health system in general [7], [39]. At the primary health care facilities, the situation has been worse despite enormous efforts that are being directed to combat the pandemic. This is because in LMICs there was a lack of preparation of primary health care governance organs responsible for making decisions to combat the pandemic [40]. The governance of the primary health care facilities is significantly needed now than ever because of the decisions that HFGCs have to make to combat COVID 19. Since primary health facilities can not be depending on decisions made by the top officially upper levels [41], HFGCs are advantageous to make contextual-based and timely decisions and institute relevant governance strategies that can minimize the COVID 19 effects at the subnational level. Strengthened governance institutions such as HFGCs promise for effective governance of COVID 19 and other pandemics in LMICs. This study has revealed some important decisions that have been made by the empowered and functional HFGCs that in turn have facilitated the operations of primary health facilities in the times of COVID 19.

## Conclusion

Effective governance of pandemics such as COVID-19 is very important for minimizing the effects of the pandemic in primary health care. strengthened governance institutions promise for achieving effective governance through applying their powers and freedom in initiating some interventions in the primary health care facilities. While there is evidence of some governance initiatives which are continuing to be taken by HFGCs as governance structures of primary health care facilities, still capacity building on the powers and governance responsibilities for these HFGCs is needed to fully explore their potentiality. The functionality of HFGCs in low and middle-income countries in times of COVID-19 is promising but still needs to be enhanced to unpack HFGCs potentials and adequately respond to the pandemic.

## Data Availability

All data produced in the present work are contained in the manuscript

## Contribution

AMK and MM were responsible for the conception of this study, research design, data collection, data analysis and writing the final draft. CM was responsible for the overall leadership of the team (setting the direction and planning) and the review of every part of the manuscript. All authors were responsible for the recruitment of research assistants and review of the final draft for submission

## Provenance and peer review

Not commissioned; externally peer-reviewed.

## Data availability statement

All data relevant to the study are included in the article or uploaded as supplementary information

